# Analysis of internet trends related to medications for COVID-19 in ten countries with the highest number of cases

**DOI:** 10.1101/2020.09.08.20190785

**Authors:** Josimar E. Chire Saire, Roselyn Lemus-Martin

## Abstract

During the last months, the pandemic generated by coronavirus SARS-CoV-2 moved governments, research groups and health organizations to plan, test and execute health policies. At the beginning, the treatment was not clear but in the next months, research groups have conducted studies to recommend or ban some medicines. At the same time, countries with most cases were changing from Asia, Europe to America. Different treatments and medications have been tested and recommended in many countries, such as Hydroxycloroquine, Ivermectin, Azithromycin, Dexamethasone, Prednisone, Remdesivir. This paper is a preliminary study about which medications people are searching on the Internet, considering ten countries with most cases in the world such as Chile, Spain, United Kingdom, Brazil, United States, India, Russia, South Africa, Peru and Mexico.

## Introduction

Since first reported in Wuhan, China, in late December 2019, the outbreak of the novel coronavirus now known as SARSCoV-2 has spread globally. As of the second week of August 2020, the novel coronavirus disease (COVID-19) pandemic has now reached over 21 million confirmed cases worldwide and approximately 762,000 deaths worldwide according to the Johns Hopkins University Coronavirus Resource Center. COVID-19 case numbers are alarming in both their volume and widening geographic scope but there are also concerns about the accuracy of reported COVID-19 case numbers, particularly at earlier stages of the pandemic and in the last months within the Latin American region, and whether under-reporting may have blurred the true extent of the outbreak from the beginning and its health and economical impact. There are many studies using Google Trends to analyze, forecast topics related to dengue (Anggraeni and Aristiani 2016), elections (Polykalas, Prezerakos, and Konidaris 2013), programming (Chen and Xing 2016) and recently, studying the COVID-19 topic in Ghana (Chire and Panford-Quainoo 2020), France (Chire and Oblitas Cruz 2020), Central American region (Chire and Lemus-Martin 2020) South American Region (Chire and Mahmood 2020).

The main contributions of this paper are:

- To study the interest of citizens through Google Search tool around medication for COVID-19
- To analyze the differences between trends in countries with high incidence of cases

## Proposal

The presented paper follows a methodology inspired on Cross Industry Standard Process for Data Mining(CRISPDM) (Shearer 2000). The steps for the conducted study are detailed in the next subsections.

### Selecting the scope of the analysis

The first question to answer rose from the announcements of the new vaccines against COVID-19, therefore the question is: what treatment or medication is most popular in the population? By consequence, to set the scope of the study, countries with the highest number of cases were selected. The analysis considers the ten countries (see Fig. 1) with highest number of infections.

**Figure 1:**
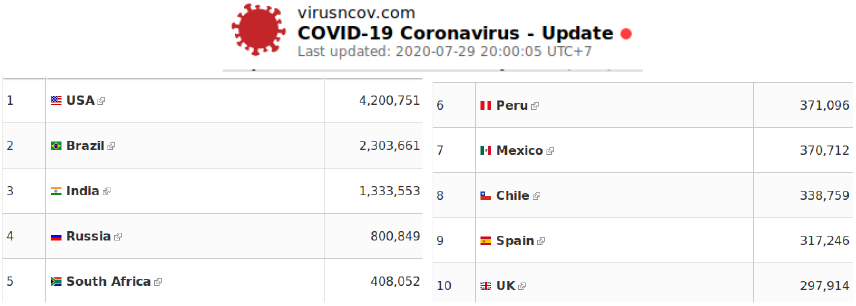
Ten countries with the highest total cases^1^

### Find the relevant terms to search

The proposed terms are related to medications used in different countries, the terms are: Chloride Dioxide, Ivermectin, Azithromycin, Dexamethasone, Prednisone, Remdesivir. These terms were selected after reviewing the news on different treatments against COVID-19 in different countries.

### Build the Query and collect data

Trends using Google search can be an useful tool to analyze how the interest of one country/city around one term is. The chosen period of study was from 01-07-2020 to 22-07-2020 and the terms were mentioned previously.

### Visualization

The collection of data from Google Trends helps to present data and support analysis about medications for COVID-19 in Top 10 countries. The authors plotted this data to show the trends evolution from 1st to 22nd July for the selected terms (figure 2)

**Figure 2:**
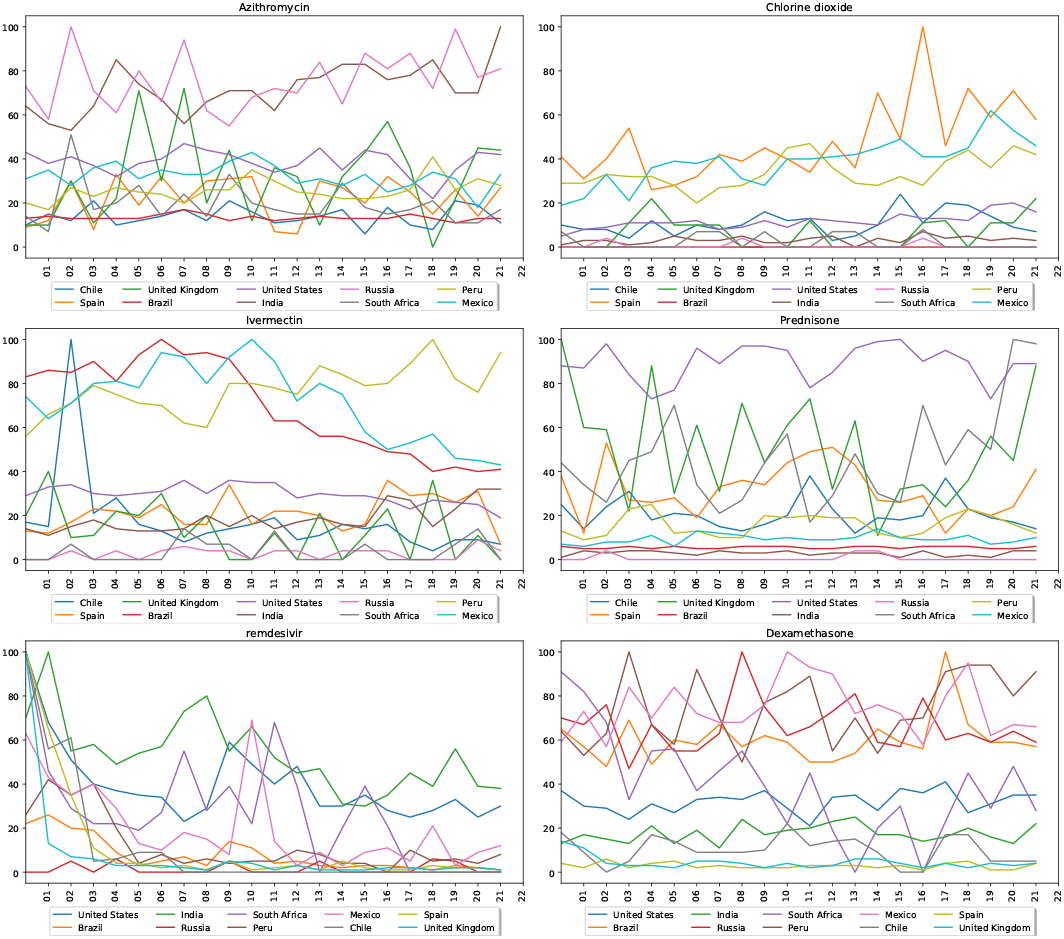
Terms: Azithromycin, Chloride Dioxide, Ivermectin, Prednisone, Dexamethasone, Remdesivir

## Results and Discussion

### Population of countries

The analysis is considering the next countries: US, Brazil, India, Russia, South Africa, Peru, Mexico, Chile, Spain, UK. Therefore, we selected countries in different regions: North America, South America, Asia and Europe. Table 1 presents the population of every country, number of COVID-19 total cases and proportion of total infections.

**Table 1:**
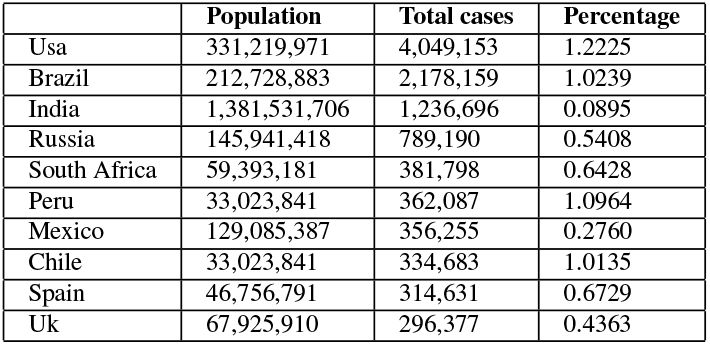
Statistics from countries

It is important to consider that each country has different population density therefore, in order to observe the total number of cases it is appropriate to analyze the proportion of total infections over the total population. If the analysis only considers data related to total cases in Table 1, then US, Brazil and India are the three nations with highest number of infections. But, considering the column Percentage in the Table 1 then US, Peru and Brazil are on the top. At the same time US is not the most populated country in the world neither Brazil or Peru.

### Google Trends Results

Performing the query using the terms mentioned previously in proposal section, the authors obtained the results shown in Figure 2.

Rusia and India present a constant interest for Azythromycin. Chloride dioxide has the highest interest in Peru, Mexico and Spain. Brazil and Mexico have a decay of interest on Ivermectine and Peru an increasing interest on Ivermectine during the last week. Prednisone has a constant interest in the United States. Remdesivir has a global decay of interest from 01 July and a constant value for India. Dexamethasone presents a constant interest in Russia, Brazil, Mexico and Peru.

In conclusion, it is possible to identify the interest of users on different COVID-19 medications using Google Trends, which considers the searches made in different countries to present the data. Besides, citizens from the countries with the highest number of infections are constantly searching information about medications which obviously shows a constant worry about the COVID-19 pandemic and opens up the issue of self-medication to get a fast alleviation and response.

## Data Availability

The data can be obtained using the parameters detailed in the paper.

## Footnotes

1 Data from https://virusncov.com/

## References

Anggraeni, W.; and Aristiani, L. 2016. Using Google Trend data in forecasting number of dengue fever cases with ARIMAX method case study: Surabaya, Indonesia. In 2016 International Conference on Information Communication Technology and Systems (ICTS), 114–118.

Chen, C.; and Xing, Z. 2016. Towards Correlating Search on Google and Asking on Stack Overflow. In 2016 IEEE 40th Annual Computer Software and Applications Conference (COMPSAC), volume 1, 83–92.

Chire, J.; and Lemus-Martin, R. 2020. Infoveillance to Analyze Covid19 Impact on Central American Population doi:https://doi.org/10.1101/2020.05.26.20113514.

Chire, J.; and Mahmood, K. 2020. Hope Amid of a Pandemic: Is Psychological Distress Alleviating in South America while Coronavirus is still on Surge?

Chire, J.; and Panford-Quainoo, K. 2020. Twitter Interaction to Analyze Covid-19 Impact in Ghana, Africa from March to July.

Chire, J. E.; and Oblitas Cruz, J. F. 2020. Study of Coronavirus Impact on Parisian Population from April to June using Twitter and Text Mining Approach. medRxiv doi:10.1101/2020.08.15.20175810. URL https://www.medrxiv.org/content/early/2020/08/18/2020.08.15.20175810.

Polykalas, S. E.; Prezerakos, G. N.; and Konidaris, A. 2013. An algorithm based on Google Trends’ data for future prediction. Case study: German elections. In IEEE International Symposium on Signal Processing and Information Technology, 000069–000073.

Shearer, C. 2000. The CRISP-DM Model: The New Blueprint for Data Mining. Journal of Data Warehousing 5(4).

